# Assessing the uptake and implementation of index testing among adolescents and young people in Sub-Saharan Africa- a systematic review

**DOI:** 10.1101/2023.09.19.23295754

**Authors:** Denise M. Sam, Clare Bukirwa, Sintieh Ekongefeyin

## Abstract

Adolescents and young people in Sub-Saharan Africa (SSA) face high mortality rates due to the Human Immunodeficiency Virus (HIV), highlighting the need for effective case identification strategies. The index testing strategy has shown promise, but its implementation and potential for use among this population remain poorly understood. We conducted a systematic review of the literature, searching multiple databases and repositories for studies conducted in SSA since 2010, involving individuals aged 10 to 24 years. Out of 1,448 studies initially identified, only three met the inclusion criteria. The implementation of the index testing strategy primarily involved recruiting index clients at healthcare facilities and testing points, followed by community-based testing. The involvement of community health workers and peer mentors proved valuable in achieving high yields. However, the legal age for consent poses a barrier to utilizing this strategy among adolescents and young people. By addressing barriers and improving facilitators, the index testing strategy holds great potential for effective case identification in this age group in SSA.

## Introduction

Over 75 million people are living with (Human Immunodeficiency Virus/ Acquired Immune Deficiency Syndrome (HIV/AIDS) globally, with a cumulative 32 million deaths [1]. With just over 12% of the global population, Sub-Saharan Africa (SSA) is home to about 71% of persons living with HIV, having ten countries accounting for 80% of cases with a prevalence of 3.6% [2,3]. This makes SSA the region with the highest HIV disease burden in the world. To address the problem of the HIV pandemic the Joint United Nations Programme on HIV/AIDS (UNAIDS) launched the 95-95-95 targets in 2014 (Heath, Levi, and Hill, 2021). The global goal of reaching 95-95-95 (95% of persons with HIV know their status, 95% of persons who know their status initiated on Antiretroviral Therapy (ART), and 95% viral suppression among persons on Antiretroviral drugs (ARV)) is key to ending AIDS as a public health crisis.

### Public health impact of HIV testing among adolescents and young people in SSA

Knowledge of an individual’s HIV status has been proven to be essential for healthy living, be it a positive or negative HIV status [5]. HIV testing is the first step in HIV prevention for those who test negative as well as a gateway to lifesaving treatment for those who test HIV positive and preventing further spread of the virus. For adolescents and young people, HIV testing proves to be more beneficial. Persons within this age group face a higher risk of HIV infection due to their rapid physical, biological and structural changes [6]. Also, young people make up part of the key populations that are disproportionately affected by HIV infection. This includes men who have sex with men (MSM), female sex workers (FSW) and injecting drug users (IDU) [7].

In 2016, over 2.1 million adolescents were living with HIV with about 84% of them in SSA [8]. Out of the over 2.7 million children living with HIV in 2022, about 1.7 million of them were between the ages of 10-19 years [9]. By 2017, about 3.9 million [2.1–5.7 million] adolescents and young people aged 15–24 were living with HIV with slightly over three out of every four of them living in SSA [10]. Even though adolescents and young people are disproportionately affected by HIV infection, HIV testing which is primordial for HIV prevention as well as linkage to treatment is still very low with just about 20% of boys as well as 30% of girls between the ages 15-19 in Africa reported having been tested for HIV [11]. Low testing rates among these age groups imply late presentation to health facilities often with advanced disease symptoms which at times leads to poor clinical outcomes [12]. As such, HIV-related mortality has not changed over the years among adolescents as compared to other age groups [13]. Improving access to HIV testing among adolescents and young people.

Achieving the 95-95-95 targets can only be successful if it can be achieved across the different subgroups. One of these is the adolescents and young people age group [14]. HIV testing is a key step in achieving HIV epidemic control, with strategies targeting this age group being a necessity. Adolescents as defined by the World Health Organization (WHO) are individuals between the ages of 10-19 years while youths are individuals within the 15-24-year age group. Young people cover the age range of 10-24 years (WHO, 2022). Out of the 1.8 billion young people in the world today, 90 per cent of them live in developing countries (Andrew Mason, 2010). According to UNICEF (2020), young people (ages 10-24) accounted for over 410,000 (194,000-690,000) new HIV infections, among whom 150,000 (44,000-310,000) were adolescents. Knowing that without treatment, adolescents and young people have a higher risk of dying, this age group, therefore, needs special attention as far as case identification and linkage to care are concerned [15]. One of the testing strategies, index testing is effective in reaching the first 95 targets [16].

Index testing which also can be referred to as partner testing/partner notification services is a testing strategy whereby contacts who have been exposed to HIV (i.e., sexually, by birth, or through sharing of needles) through contact with an HIV-positive person (considered the index client), are identified and offered HIV testing services [17]. The index testing strategy has been proven in several studies to be very effective in the identification of new HIV cases which is a necessary step in improving access to HIV care for those infected as well as HIV prevention for those tested negative [18–20]

Several strategies have been used over the last decade with varying efficacy to increase testing uptake among adolescents and young persons in SSA. Index testing is one of these strategies that has been proven to have one of the highest uptakes and yields in terms of positivity among those tested [21,22]. As a targeted testing strategy, it is also considered more cost-effective and sustainable compared to voluntary testing and counselling strategies in terms of the number of tests carried out and the testing yield [23].

Noting its high efficiency in the identification of new HIV cases, the WHO recommends Index testing as one of the main testing strategies as part of a package of testing services, with adolescents being one of the target groups [24] According to the WHO, several countries have adopted and recommended the Index testing strategy, however, less than half of them show evidence of actually implementing the strategy [24]

This systematic review aims to identify barriers, facilitators, challenges, and opportunities in the implementation of index testing among adolescents and young people in SSA. The study seeks to improve the effectiveness of the strategy and bridge the testing gap in the region. The research questions are as follows:

1. How is index testing implemented among adolescents and young people in SSA?
2. What are the barriers and facilitators to index testing in this population?
3. What challenges and opportunities are associated with implementing the index testing strategy among adolescents and young people in SSA?

## 3. Methods

### Study design

To answer the research questions, a systematic review was conducted. The choice of a systematic review instead of primary research stems from the need to provide a more comprehensive view of the subject. This will avoid a situation where only one particular country’s experience in the region is considered. Secondly, the systematic review had the possibility of including primary studies carried out using different methodological approaches such as programmatic implementation and cross-sectional studies to answer the research question.

This systematic review was carried out following the Preferred Reporting Items for Systematic Reviews and Meta-Analyses (PRISMA) recommendations [25]. A systematic review proposal was written and submitted to the University of Edinburgh. This proposal was registered with Prospero (Registration number: **CRD42022362682**) to avoid any biased changes in methodology during the search process.

### Ethical Considerations

To ensure that this study was ethical, an application to assess ethical integrity and risk was submitted to the Usher Masters Research Ethics Group (UMREG) of the University of Edinburgh. Data used in this review were publicly available data with no access to data which counld identify the participants in the primary studies. As such, there was no need for informed consent or parental consent. Following the completion of the Research Ethics Self-Evaluation and Review Form (RESERF), the study was considered as posing no reasonably foreseeable ethical risks and therefore considered to be ‘exempt’ without the need for further ethical review..

### Search Strategy

To support the whole review process Covidence was used. Covidence (http://www.covidence.org/) is a web-based tool used for data screening and extraction and it is one of the tools recommended by the Cochrane collaboration [26]. The tool permits the use by multiple reviewers to simultaneously screen articles, carry out full text reads, customise and evaluate the quality of included papers as well as identify and remove duplicates.

The PICOS (Problem/Population, Intervention, Comparison Group, Outcome(s), and Setting), adapted from Syrene A. Miller, PICO Worksheet and Search Strategy National Center for Dental Hygiene Research [27] criteria as seen below was used to develop the search terms used. After discussing with colleagues, alternative terms were identified as seen in Table 1

**Table 1.** PICO (Problem/Population, Intervention, Comparison Group, Outcome(s), and Setting)

| PICO | Terms | Alternate terms |
| --- | --- | --- |
| Problem/Population | Adolescents (10-19 years) and<br>Young Adults (10-24years) | Adolescents/Young Adults<br>10-19, and 20-24years<br>Adolescents<br>Young persons |
| **Intervention** | Index testing (contact tracing, partner notification services) implementation | Index testing<br>Partner notification service<br>Contact tracing<br>Family testing<br>Index Case Testing |
| **Comparison** | None |  |
| **Outcomes** | HIV testing uptake and Implementation |  |
| **Setting** | SSA | West Africa, Central Africa, Eastern Africa, Southern Africa |

### Grey literature

The WHO institutional repository [28] as well as the repository for the International AIDS society [29] were searched for any articles, abstracts or papers that met the inclusion criteria. Secondly, the reviewers searched online for National strategic plans for HIV as well as guidelines for the management of HIV for countries in SSA. Any papers found were uploaded to Covidence for screening and full-text reading.

Note that, any disagreements or differences were settled by ES Or have a third opinion.

S1 Tables show the search terms used. Three main electronic databases were searched systematically for peer review publications; PubMed, Cochrane Library and EMBASE. These databases were chosen given that together they contain a significant number of published articles [30,31]. Secondly, these databases contain publications that are relevant to answering the research questions.

### Study Eligibility and selection

The two reviewers independently screened through the search results and selected studies and articles that met the above eligibility criteria. The reviewers began by carrying out a preliminary title and abstract screening in the various databases. Studies that met the eligibility criteria were then downloaded and saved in the reviewers’ computers before being uploaded to Covidence.

After uploading all the articles into Covidence, duplicates were identified and removed. A second title and abstract screening were carried out by the two reviewers in Covidence followed by a full test review of the included articles to ensure they met the inclusion criteria.

### Quality Assessment

To ensure that the studies included were of good quality, the Critical Appraisal Skills Programme (CASP) checklist was used [32]. This tool was selected because it is more commonly used in the quality appraisal of health-related qualitative research as well as its endorsement by the Cochrane Qualitative and Implementation Methods Group [33]. The data collection sheet in Covidence was customised as well as the quality appraisal forms. This permitted the reviewers to assess the quality of different study types included in the review. To avoid bias, two reviewers were independently involved in the assessment of the quality of the included studies. Any disagreement was resolved by ES.

In total, there were 13 questions in the customised assessment tool, however, not all studies were subjected to all 13 questions. For each question, the responses were, “YES”, “NO”, “UNCLEAR” and “NOT APPLICABLE”. Given that the website (https://casp-uk.net/faqs/) did not recommend any particular score for the tool. However, it proposed a quality grading the studies as “poor”, “moderate”, and “high”. We therefore used this grading system to assess the various studies. Having a score of <50% was considered low quality, 50% to <100% as moderate quality while a score of 100% was considered high quality.

### Data Extraction

The following data were extracted from the selected articles: the author and year of the study, where the study was carried out, study design, sample size and response rate, the testing strategy used in the study, the testing location as well as who was in charge of carrying out the test. The other information extracted from the studies included facilitators of uptake among index and contact persons, barriers to uptake among index and contact persons, challenges and opportunities of implementing index testing and finally, the strengths and limitations of the study.

### Data Analysis and synthesis

A qualitative analysis and narrative synthesis were carried out with a focus on facilitators and barriers of the uptake among the index persons and the contact persons, the challenges and opportunities implementing the strategy as well as any percentage uptake and yield of the index testing. Meta-analysis could not be carried out given the heterogeneity of the included articles. Articles and grey literature retrieved from databases were exported to Mendeley Reference Manager version 2.77.0.

## 4. Results

### Search Results

Following a search of the various databases, 1,451 studies were identified, including 1,343 studies from PubMed, 9 studies from Embase, 96 studies from the Cochrane database, and 3 studies from other sources. The initial screening of the 1,448 studies was done by this author, the primary reviewer. This was a review of the titles to ensure that they met the inclusion criteria. A total of 1,284 studies that did not meet the inclusion criteria were not selected. Following this exclusion, 167 studies were then selected and uploaded to Covidence which automatically identified 15 duplicates which were removed.

At this stage, both reviewers were involved in the review process. Each reviewer screened titles and abstracts of 152 studies resulting in the exclusion of 122 studies. The primary reviewer identified studies without full text and searched the internet and the University library to get them. The full texts for 30 studies were uploaded to Covidence. The two reviewers then proceeded to assess for eligibility and quality, which led to the exclusion of 27 studies with reasons noted. Fig 1 summarises the various steps in the review process.

**Figure 1.**
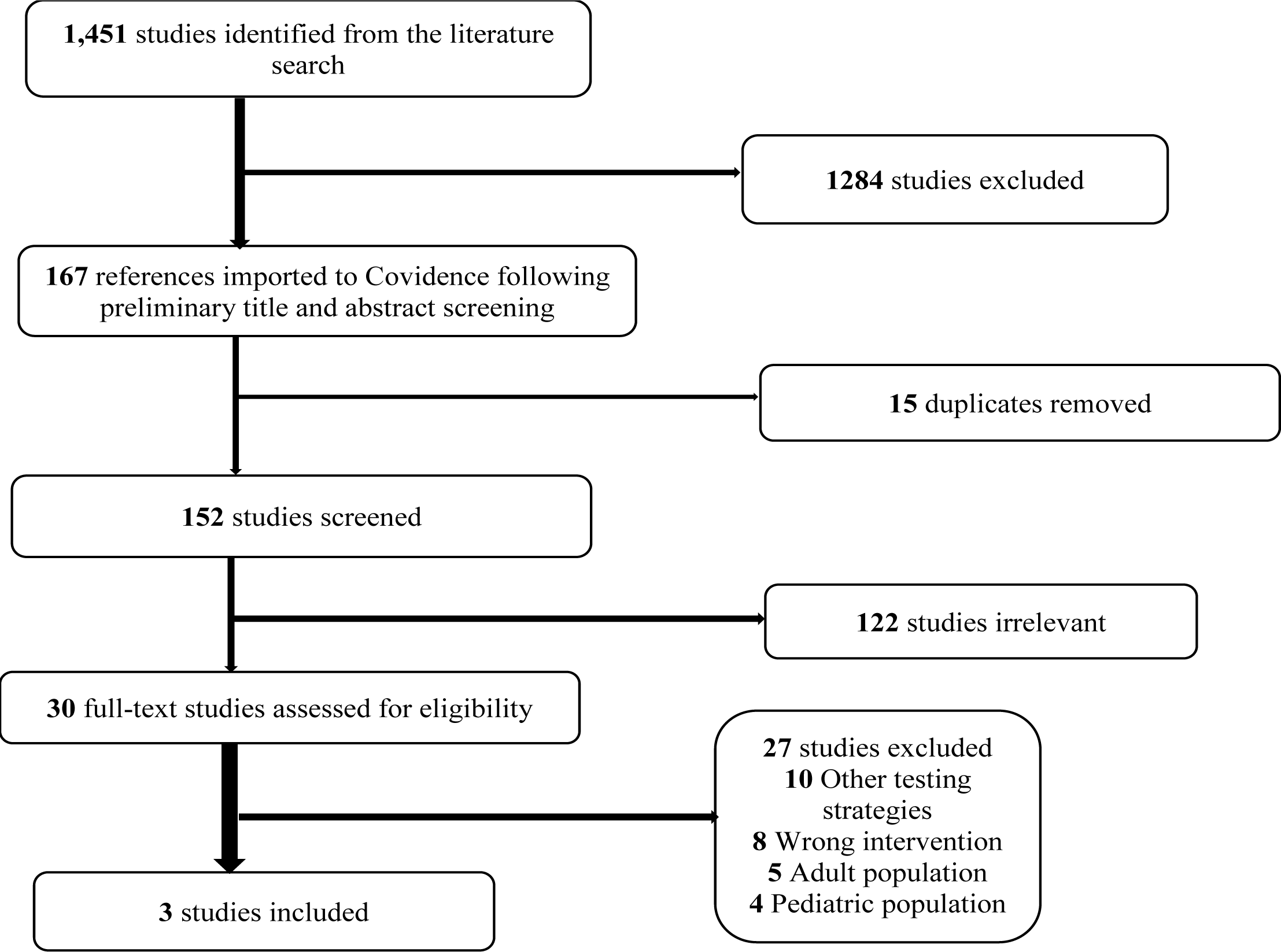
PRISMA Flow chart for the study selection *(adapted from the PRISMA diagram)* [25].

The search for grey literature resulted in three abstracts from WHO institutional repository and the International AIDS Society conference abstract database that were included in the total studies identified (n=1,451). However, these studies did not meet the eligibility criteria for full-text read and so were not included in the final analysis. As such, all three included studies were peer-reviewed.

### Study Characteristics

Following a full-text assessment, three studies were included in the review[20,34,35]. S3 tables represent the different characteristics of the included studies.

### Quality assessment

The three included studies were assessed for quality before extracting the data. A summary of the CASP analysis for quality can be found in S1 Table. In this review, all included studies were evaluated to be of moderate quality

As seen in the S3 tables, these three studies were carried out as part of the implementation of HIV testing programs that took place between 2014 and 2018. While two of the three studies had their data collected during implementation, one of them used programmatic data after the implementation process was completed [35]. Two of the three studies involved study participants between 0 and 24 years [20,35] while the other had study participants aged between 15 and 19 years old [34]. Only one of the three studies disaggregated the 15-24 age group by gender with 40.6% (n=69) being male and 56.4% (n=93) females [20].

The total sample size for the three studies was 23,666 participants, however, the sample size of participants within the desired age group for this review (10 to 24 years) was 11,167 participants.

### Index testing implementation strategy

The three studies included in the review implemented the index testing strategy with some differences in terms of recruitment and testing location for the contacts.

All three studies identified Index persons at the health facilities with follow-up of the contact persons done at different locations, however, one study also recruited index persons at mobile unit testing points as well as door-to-door testing [34]. Jubilee et al. carried out community testing of contact persons while Ahmed *et al.* and Tapera *et al.* gave options to the index persons to choose between facility and community or home testing. In addition, Tapera *et al.* also gave a third option of HIV self-testing to persons who didn’t want any of the other options.

The age of consent in Jubilee *et al.* was 12 years while for Ahmed *et al.* was 16 years. Community health workers were in charge of carrying out testing in two of the studies [20,35] while HIV testing and counselling (HTC) counsellors were in charge of testing in one [34]. See Table 7 for more on the index testing implementation process. While Ahmed *et al.* considered all household members of the index persons as contacts for testing, Tapera *et al.* were more interested in households with members less than 25 years old and Jubilee *et al.* targeted mainly, sexual partners and biological children.

### Facilitators and barriers to Index testing

S1 Table 3 summarises the facilitators, barriers, challenges and opportunities linked to the index testing strategy while Fig 2 illustrates the interconnection.

**Figure 2.**
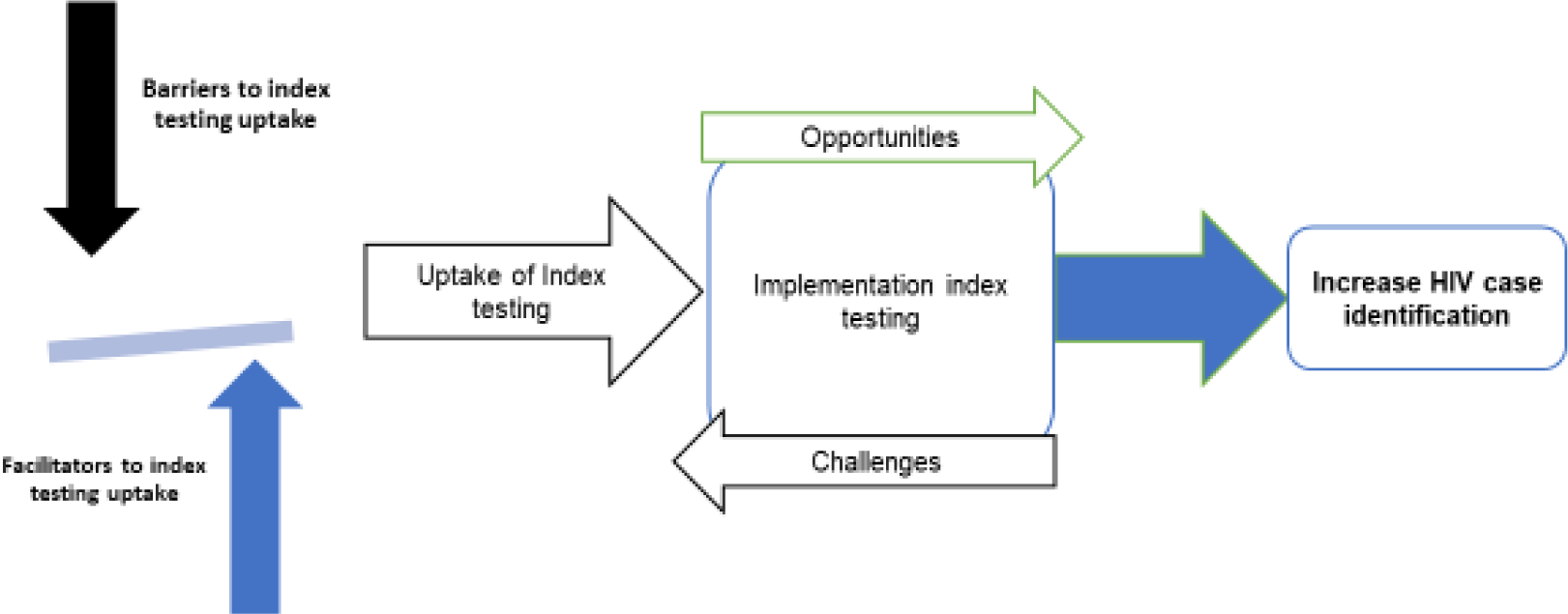
Connection between the factors associated with Index testing.

### Facilitators

Tapera *et al.* had an overall index testing uptake of 85.17% with an uptake of 83.7% among participants aged between 15-24years old, which was far better than the national average of 50.4% of 15-24-year olds who knew their HIV status. The overall yield (both adults and young people) of testing was also high with 9.8% of all those tested individuals resulting in a positive HIV test result (1,193 positives out of 12,114 tested). The use of an adolescent peer-led model known as community adolescent treatment supporters (CATS), which involved training using a well-structured curriculum as well as on-the-job mentorship was found to be useful in achieving the high output in the program. This model was not only useful in the testing process but was also primordial in achieving great linkage and treatment outcomes [35].

Ahmed *et al.* also found a high uptake of index testing, with 93.5% of persons consenting to the strategy with a majority (88%) preferring home testing over facility testing. The yield among the 15-24year olds was 4.2% (7 positives out of 165 tested). Home testing was therefore found to be instrumental in meeting young persons who may not be found around health facilities [20].

As for Jubilee *et al.,* out of the 7,916 persons approached for index testing at their homes, 5,937 (75%) accepted with a final overall testing yield of 4.2% (454 positives out of 10,854 contacts tested). In this study, the positivity rate among adolescents aged 15-19 was 2.4% per cent, however, the positivity was significantly higher among adolescents who were identified as sexual partners with a yield of 22.6% (7/31) unlike those who were biological children to the index persons with a yield of 1.8% (19/1,057). The study identified offering index testing to newly diagnosed HIV-positive individuals as a key factor in achieving high yields. After further analysis, the study also reported that persons tested using the index testing strategy were more likely to get linked to care than those tested through other strategies [34].

### Barriers

Tapera *et al.* found very low testing uptake among lower age groups such as those below 9 years as compared to those between 20 and 24 years given that parents need to be available to provide consent for these participants to be tested [35]. The other two studies did not mention specific barriers in this case.

### Challenges and opportunities with implementation of Index testing among adolescents and young people

#### Challenges

A common challenge identified by Jubilee *et al.* and Ahmed *et al.* was that even though most participants preferred home testing to facility testing, there was difficulty in meeting participants at home during these scheduled home visits [20,34]. Tapera *et al.* also observed that even though the participants had an option for self-testing and home-based testing, the uptake was lower in their study. This finding was unlike that observed by Jubilee *et al.* who found home-based testing as a more acceptable testing option.

#### 4.5.2 Opportunities

Fig 2 shows the relationship between the various factors affecting the uptake and outcome of the index testing implementation process. Jubilee *et al.* observed a higher positivity rate among first-time testers compared to re-testers who were offered index testing as well as a higher yield among contacts of newly diagnosed index persons compared to contact persons of index persons already on treatment [34]

## 5. Discussion

### Implementation of index testing among adolescents and young people in Africa

The three studies presented above describe the implementation of the index testing process among adolescents and young people in three SSA countries. The studies describe an implementation process that begins with the identification of index persons mostly in health facilities but also at HIV testing points such as mobile testing units and door-to-door testing. This was followed by seeking the consent of the index persons to test members of their households as well as sexual partners. This showed varying uptakes in the different studies ranging from 75% to 93.5%. This can be considered acceptable especially considering the 95-95-95 targets.

The authors highlight the importance of peer mentors and community health workers (CHWs) in the delivery of index testing. This adds to the growing evidence of the importance of CHWs in the different aspects of HIV programming [36–38]. This, therefore, may imply working with CHWs might be useful for achieving index testing among adolescents and young people given that they supplement the health workforce to carry out services even at the community level, meeting young people who may not visit health facilities.

Using peers to improve testing among adolescents and young people is also a recommended option for improving testing among this age group [39].

Even though a recommended approach to testing, many countries are yet to fully utilise peer mentors in their testing programs. Further research may be needed to further understand the use of using peer mentors and community health workers in the HIV testing process.

The next step in the implementation was testing the contacts given by the index person. The index persons were offered the options of visiting their homes to test their family members or referring the contact persons to health facilities to be tested as well as taking the self-testing option. These options are in line with previous recommendations for alternate HIV testing service deliveries for adolescents and young people (ibid). Even though there were no specific tailored centres for adolescents and young people, Tapera *et al.* involved young peer mentors who carried out other treatment support as well as education activities geared towards adolescents and their caregivers. This could be an important lesson for other program implementers carrying out similar programs as adolescents and young people tend to trust their peers more, during testing [40].

The implementation of the index testing among adolescents and young people reported in these studies had a slightly different approach to index testing among adults in the area of partner notification. Among adults, it often involves notification of contacts either by the index persons or the health care personnel. However, this, was not often the case as permission to come to test the household was taken from the index persons. This highlights another peculiarity with index testing among young persons which could be a limitation to getting adolescents and young people tested using this strategy.

### Barriers and facilitators of index testing among adolescents and young persons

One of the main barriers identified in the studies was the legal age of consent for testing which affected uptake among adolescents below certain age groups. This finding corroborates previous studies identifying the legal age of consent to HIV testing as a barrier to HIV testing uptake among adolescents and young persons. This high legal age of consent for HIV testing implies that young persons will need parental permission to get tested which may dissuade them. Unfortunately, many countries in SSA have not considered this in their national testing guidelines [41,41]. The identified studies did not focus on identifying the barriers to uptake, however, other studies have identified barriers such as fear of stigma, fear of having positive results, limited access to HIV testing, and non-disclosure of HIV status by parents and partners [42–44]. This may have been reflected in the implementation given that not all contacts were home during the testing by the CWHs.

In the three studies, different strategies showed to help improve the uptake of the index testing process. These included the use of the adolescent peer-led model, providing differentiated service delivery, that is services targeting specific needs to different subgroups as well as offering several testing options including home testing. These strategies are similar to those proposed by other studies that identified education and addressing stigma fears to be facilitators for HIV testing[45,46], testing adolescents at a location where they feel more comfortable could reduce the fears, thereby facilitating the uptake of HIV testing. Knowledge of these barriers and facilitators can be useful in better designing index testing programs for adolescents and young people that will be more accepted thereby increasing uptake of the strategy.

### Challenges and Opportunities of the index testing strategy

Index testing as a strategy faces certain challenges but also has new opportunities as an HIV testing strategy. Even though contacts preferred home testing, the main difficulty is getting to meet them during visits by the CHWs. This included parents not being available to give consent for the testing of their children. This reduces the number of adolescents who can get tested at the community level and at times links the testing of adolescents to the willingness and availability of the parent which in some cases may lead to a missed diagnosis. Another identified challenge was the low uptake of HIV self-testing as a testing option during index testing [35]. Even though not described in the study, a previous study identified a lack of status disclosure as a reason for the low uptake of self-testing during index testing [47]. Another challenge in the study was the fact that most adolescents were tested within a household testing context which may lead to coercion and deprive them of their autonomy as recommended by the WHO [43]. Other challenges like the risk of social harm like stigma, and discrimination resulting from testing were not considered in the studies within this review [43].

Even though faced with many implementation challenges, these studies all showed high positivity compared with other testing strategies like the door-to-door and fixed-point testing reported by Jubilee et al. (2019). This makes it a sure strategy to achieve epidemic control among the adolescent and youth populations. As such, program implementers can take these opportunities and challenges into account when designing index testing programs for adolescents and young people. This will reduce difficulties faced during implementation while capitalising on the opportunities.

### Strengths and limitations of the review

#### Strengths

This review included studies with a significant sample size in three different countries within the SSA region. Secondly, this systematic review made use of evidence from routine programmatic implementation of the index testing strategy among adolescents which showed some realities on the field and not just in research. Therefore, this study presents findings that can be very helpful for programmatic implementation. In addition, it combines evidence from three different countries in SSA, making it an opportunity to gain a broader experience of the region other than a single country experience.

#### Limitations

The review had several limitations. The studies included were all carried out as part of a programmatic evaluation with one study retrospectively using program data. This limited the study type and may have been the source of bias. This is because most funded programs during implementation may follow certain donor specifications and guidelines which may introduce bias in the coverage and outcome indicators. In addition, all three studies focused on the implementation process and did not evaluate factors associated with certain outcomes. Also, the review of grey literature did not yield any additional studies to be included. Even though websites of certain national HIV programs were searched, only a few of them had their HIV guidelines included and among those who did, very little was mentioned concerning index testing among adolescents and young persons. The few documents identified from the grey literature did not meet the inclusion criteria.

### Public health implication

HIV infection is a public health crisis on a global scale, that affects different subpopulations in different ways and to successfully achieve epidemic control, there is a need to have a multilevel approach to addressing the pandemic. Index testing has been shown in this review and from previous studies to be effective in the identification of new HIV cases, both in the community and in the health facility. It presents itself as an innovative strategy in helping adolescents and young people know their status, an important phase in helping those living with HIV to begin life-saving ART. Even though useful and effective in case identification, this review has highlighted some factors on which adolescents, caregivers, policymakers, and society can either capitalise on or improve to benefit from the testing strategy.

Within communities in SSA, there is a need for more communication to improve the awareness of HIV testing among adolescents and young people. This includes disclosure of HIV status by parents, guardians and partners living with HIV, as well as communication at the community level to reduce the stigma around HIV testing in general within this age group. There is a need to create more adolescent and youth-friendly centres that can encourage persons within these age groups to test without being stigmatised.

Index testing among adolescents and young people can benefit more from the use of peer mentors and community health workers to assist in the uptake and testing process. There is a need for countries within the SSA region to begin adopting policies to better make use of these CHWs in addressing the HIV pandemic in general as well as index testing among adolescents and young persons. One of the most important points to consider is to listen to the voices of adolescents and young people in every program concerning them. No one can decide what works best for adolescents and young people other than they do. This includes putting in place legislation that respects the rights of adolescents and young people, giving them the right to have access to HIV testing as well as the right to choose whether or not they want to get tested. Furthermore, the needs of adolescent girls and young women are different from those of adolescent boys and young men. This implies that gender equality needs to be a major factor, giving both boys and girls an opportunity to have their voices heard.

With the recent rise in recognition of the importance of CHWs in achieving health-related sustainable development goals [48], it is no doubt that making them part of the health system will significantly improve health outcomes. There is also a need to encourage countries to address laws targeting the age of consent to permit more adolescents to access HIV testing services as a whole and index testing in particular. Another important recommendation from this review is for national HIV programs to include sections focused on index testing among adolescents and also make available these guidelines public to ensure health providers get access to these documents to permit them fully implement.

This review has identified that most index testing programs are mostly funded as part of routine HIV prevention, care and treatment programs from major donors. However, there is a need for these donors to not only fund routine programs but also implementation studies to better understand how strategies such as index testing can be more effective. These studies can better shed light on these strategies, helping policymakers at the country level to easily adopt and implement the strategy. More funding needs to go to researchers understanding and improving upon the strategy to achieve better yields and impact in HIV case identification among adolescents and young people. Globally, there is a need to increase the visibility of research work from the SSA region. Given that very few journals are existing within the region, encouraging the publication of research work as well as lessons learned during implementation will increase publicly available data that can be used to share experiences.

### Critical assessment of the subject

The paucity of data on index testing amongst adolescents and young people may be due to one of two reasons. Either, very few of the many programs find it important to publish the work they do or index testing among adolescents and young people is yet to be as developed as for adults. Even with a grey literature search, none of the identified studies met the inclusion criteria to be included among the reviewed studies. The websites of different national AIDS control programs in SSA were searched but very few had their national guidelines available online.

There is also a need to carry out more implementation science research studies to improve the quality of these programs to improve the practicality of the strategy. Also, funders may decide to make available extra funds for implementation research or make the publication of findings a part of the project deliverables to increase the number of publicly available data.

## 6. Conclusion

This review adds to the evidence that index testing could be a great strategy to achieve optimal HIV case identification among adolescents and young people. There are still challenges with addressing the legal age of consent among this age group, which affects the uptake and implementation of the strategy. Due to the limited research, there is a need for more research such as implementation science research to better understand the operational challenges of an index testing strategy among adolescents and young people as well as to understand better ways of implementing the strategy. For index testing to be optimal, it should not only focus on case identification but also pay attention to the needs and challenges faced by the adolescents and young people who are impacted by the strategy. The strategy needs to evolve to address the stigma faced by these persons and empower them to decide when to say “Yes” or “No” to testing. Addressing the barriers and challenges will make the strategies more acceptable, but understanding the needs of the adolescents and young people in SSA vis-à-vis index testing will make it successful in achieving epidemic control of HIV within the adolescent and youth subpopulations.

## Supporting information

S1 File_Search Strategy

S2 File_Quality Assessment of included studies

S3 File_Characteristics of Evidence

S4 PRISMA checklist

## Data Availability

The authors confirm that the data supporting the findings of this study are available within the article and its supplementary materials

## Acknowledgements

We wish to acknowledge the immense and invaluable contribution of Dr Zhong Eric Chen (Senior Researcher, Faculty of Sexual and Reproductive Healthcare Clinical Effectiveness Unit, NHS Lothian, Scotland; Honorary Fellow, University of Edinburgh) and Dr Magda Mekky (Teaching Fellow for the online Master of Public Health Programme, University of Edinburg) in supervising this work.

## Supporting information

S1 File Search strings

S2 File Quality Assessment of included studies

S3 Study Characteristics

## Notes

### Competing Interest Statement

The authors have declared no competing interest.

### Funding Statement

The authors received no specific funding for this work

