## Supplementary material for "Assessing the uptake and implementation of index testing among adolescents and young people in Sub-Saharan Africa- a systematic review": S1 File_Search Strategy

S2 File Quality Assessment of included studies using the Critical Appraisal Skills Programme (CASP) checklist

Table 1; Quality Assessment of Included Studies

| Checklist questions | Jubilee 2019 | Ahmed 2017 | Tapera 2019 |
|---|---|---|---|
| 1. Were the criteria for inclusion in the sample clearly defined? | Yes | Yes | Yes |
| 2. Were the study subjects and the setting described in detail? | Yes | Yes | Yes |
| 3. Was the exposure measured validly and reliably? | Not applicable | Yes | Yes |
| 4. Were objective, standard criteria used for measurement of the condition? | Yes | Yes | Yes |
| 5. Were confounding factors identified? | No | No | No |
| 6. Were strategies to deal with confounding factors stated? | Yes | Yes | Unclear |
| 7. Were the outcomes measured in a valid and reliable way? | Yes | Yes | Yes |
| 8. Was appropriate statistical analysis used? | Yes | Yes | Yes |
| 9. Is the research ethical according to current criteria or, for recent studies, and is there evidence of ethical approval by an appropriate body? | Yes | Yes | Yes |

| | | | |
|---|---|---|---|
| 10. Do the conclusions drawn in the research report flow from the analysis, or interpretation, of the data? | Yes | Yes | Yes |
| 11. Were the specific directives for new research appropriate? | Not applicable | Not applicable | Not applicable |
| 12. Was appropriate statistical analysis used? | Yes | Not applicable | Not applicable |
| 13. Was the trial design appropriate, and were there any deviations from the standard RCT design (individual randomisation, parallel groups) accounted for in the conduct and analysis of the trial? | Not applicable | Not applicable | Not applicable |
| Total | 9/10 | 9/10 | 8/10 |
