## Supplementary material for "Assessing the uptake and implementation of index testing among adolescents and young people in Sub-Saharan Africa- a systematic review": S2 File_Quality Assessment of included studies

S3 Characteristics of Evidence

Table 1; Study characteristics

| **Author (Year)** | Jubilee M et al (2019) | S Ahmed et al (2017) | Tapera et al (2019) |
|---|---|---|---|
| **Place of study** | Lesotho | Malawi | Zimbabwe |
| **Study design** | Other: Implementation study | Other: Implementation study | Cohort study |
| **Sample size and response rate** | 1088 adolescents (15 to 19 years)<br>7982 children (2 to 14 years) | 461 for children and young persons, 165 young persons | 15,223 for 0 to 24-year-olds, 9,914 for 10 to 24-year-olds |
| **Study period** | May 2015 to Nov 2017 | July 2014 to April 2015 | Oct 2017 to Sep 2018 (data collected)<br>Jan 2019 to May 2019 (study done) |
| **Possible conflicts of interest for study authors** | Not mentioned | None | None |
| **Population Age group** | 15 to 19 | 1 to 24 years | 0 to 25 years |

| | | | |
|---|---|---|---|
| **Inclusion criteria** | All PLHIV in the identified health facilities and PSI HTS* sites, who consented to participation | - All people enrolled in the HIV treatment programme at Mponela health centre between July 2014 and February 2015<br><br>- Those who consented<br><br>- Had a household member with an unknown HIV status | Household and sexual contacts of HIV-positive children and adolescents aged 0 to 25 years. |
| **Exclusion criteria** | Not stated | If the index case lived alone or all household members had a known HIV status | Household and sexual contacts with known HIV status |

*Population Services International (PSI), HIV Testing Services (HTS)

Table 2; Index testing implementation process

| Author (Year) | Jubilee M et al (2019) | S Ahmed et al (2017) | Tapera et al (2019) |
|---|---|---|---|
| Place of study | Lesotho | Malawi | Zimbabwe |
| Testing strategy | Index testing | Index testing | Index testing |
| Location of testing | Community | Health Facility and Community | Health Facility and Community |
| HIV tests carried out by | Trained provider | Community health worker | Trained provider, self-testing, and Community health workers |
| Limitations of study | • Implemented in only high prevalence settings with more resources, findings may be different in low burden settings with limited resources. | • Underreporting of the number of family members in need of testing could have led to a response bias<br>• The study did not indicate sexual and biological contacts, they instead mentioned relationship statuses like a spouse, biological child, biological sibling, or other (without stating clearly what other meant)<br>• Instead of focusing on the transmission route, the program focused on household members, which may have led to a lower positivity yield | • Used routine secondary data which may have errors including some missing data<br>• No information on other sociodemographic and clinical data which may be associated with non-testing<br>• Other confounding factors and variables not analysed |
| Strength of study | Included large numbers of adolescents and children | Evaluated both identification and linkage outcomes of index testing | • Covered 24 out of the 63 health districts in Zimbabwe, so representative<br>• Studied a large number of contacts<br>• provision of HIV preventive services to HIV-negative |

| | | | |
|---|---|---|---|
| | | | contacts, including voluntary medical male circumcision, family planning, and cervical cancer screening for young women |

Table 3; Factors associated with the implementation of index testing

| **Author (Year)** | Jubilee M et al (2019) | S Ahmed et al (2017) | Tapera et al (2019) |
|---|---|---|---|
| **Place of study** | Lesotho | Other: Malawi | Other: Zimbabwe |
| **Study design** | Program Implementation | Program Implementation | Program implementation |
| **Facilitators for uptake among index persons** | | • The uptake of the index testing was 93.5%<br>• The majority of participants preferred home-based testing (88.2%) over facility-based testing. 156 preferred | The use of peers (Community Adolescent Treatment Supporter or CATS) |

| | | | |
|---|---|---|---|
| | | home-based while 9 preferred facility testing | |
| **Barriers to uptake among index persons** | | • Having family members and partners with known HIV status<br>• Unavailability of parents to provide consent | |
| **Facilitators of uptake among contact persons** | | | Provision of home testing for those hesitant to visit the health facility |
| **Barriers to uptake among contact person** | | • Possibility of underreporting of household members by the index persons<br>• Non-availability of participants at home during visits by community health workers | • Legal barriers requiring the consent of parents for minors<br>• hesitancy to come to the health facility for testing |
| **Challenges in implementing Index testing** | | | • Out of 9914 adolescents and young people referred 1470 refused to test. No particular reasons given<br>• Low uptake of self-testing and home testing |
| **Opportunities for index testing uptake** | | • High yield 7 positive cases out of 165 tested, 3 of whom were previously tested HIV negative<br>• Possibility of reaching young people who are not based in facility-based settings | • Uptake of 85.17%<br>• The overall positivity rate of 9.8% (including children) makes it an effective testing strategy<br>• 96.6% initiation to ART |
