## Supplementary material for "Assessing the uptake and implementation of index testing among adolescents and young people in Sub-Saharan Africa- a systematic review": S4 PRISMA checklist

S1 File: Search Strategy

Key terms :

(((Index testing) OR (Index Case testing) OR (Partner testing) OR (Partner notification testing) OR (Contact tracing) OR (Family testing)) AND ((Adolescent) OR (Young people) OR (Young persons) OR (Young Adults)) AND ((HIV testing uptake) OR (HIV test) OR (HIV test implementation)) AND ((SSA) OR (West Africa) OR Central Africa) OR (Southern Africa)))

Table 1; Search Results from Cochrane database search (23/10/2022)

96 studies found

| | |
|---|---|
| Search Name: Index testing in adolescents | Last Saved: 23/10/2022 17:43:55 |
| Search Results | 96 studies |
| #1 | index testing OR Index case testing OR partner testing OR partner notification testing OR family testing |

| | |
|---|---|
| #2 | adolescence OR young persons OR young adults OR young people |
| #3 | HIV test OR HIV test implementation OR HIV testing uptake |
| #4 | SSA OR West Africa OR East Africa OR Central Africa OR Southern Africa |
| #1AND#2AND#3AND#4 | 96 studies |

Table 1; EMBASE Search results (23/10/2022)

6 studies found

| No. | Query | Results |
|---|---|---|
| **6** | **#5** | **#1** AND **#2** AND **#3** AND **#4** |
| **495,961** | **#4** | ((('**africa south of the sahara**'/exp OR '**africa south of the sahara**' OR **subsaharan**) AND ('**africa**'/exp OR **africa**) OR '**central africa**'/exp OR '**central africa**' OR **east**) AND ('**africa**'/exp OR **africa**) OR **west**) AND ('**africa**'/exp OR **africa**) OR '**south africa**'/exp OR '**south africa**' |
| **4,170** | **#3** | ((('**hiv**'/exp OR **hiv**) AND **test** OR '**hiv**'/exp OR **hiv**) AND **testing** AND **uptake** OR '**hiv**'/exp OR **hiv**) AND **test** AND **implementation** |
| **131,801** | **#2** | ((('**adolescent**'/exp OR **adolescent** OR **young**) AND ('**persons**'/exp OR **persons**) OR **young**) AND ('**adults**'/exp OR **adults**) OR **young**) AND **people** |
| **80,957** | **#1** | (((((**index** AND **case** AND **testing** OR **index**) |

| | | |
|---|---|---|
| | | AND **testing** OR **partner**) AND **testing** OR **partner**)<br>AND **notification** AND **testing** OR **contact**)<br>AND **tracing** OR **'family'**/exp OR **family**) AND **testing** |

Table 2; PubMed Search Results (23/10/2022)

1,343 studies found

| Search query: ((((((((((index testing) OR (index case testing)) OR (partner testing)) OR (partner notification testing)) OR (family testing)) OR (contact tracing))) AND (((((adolescent) OR (young persons)) OR (young people)) OR (young adults)))) AND ((((HIV test) OR (HIV testing uptake)) OR (HIV test implementation)))) AND ((((((Subsaharan Africa) OR (West Africa)) OR (Central Africa)) OR (East Africa)) OR (Southern Africa)))) AND (2011:2022[pdat])) | |
|---|---|
| Year | Count |
| 2022 | 73 |
| 2021 | 130 |
| 2020 | 154 |
| 2019 | 157 |
| 2018 | 123 |

| | |
|---|---|
| 2017 | 111 |
| 2016 | 110 |
| 2015 | 121 |
| 2014 | 108 |
| 2013 | 101 |
| 2012 | 85 |
| 2011 | 70 |
